# High initiation but sub-optimal completion of tuberculosis preventive treatment among people living with HIV on ART, persistent tuberculosis incidence and programmatic implications: Findings from a two-year multi-country cohort study

**DOI:** 10.64898/2026.01.16.26344254

**Authors:** L Chimoyi, S Ginindza, BAS Nonyane, C Mulder, A Bedru, N Kawaza, JE Golub, K Shearer, A Kennar, L Mvusi, K Vilakazi-Nhlapo, T Apollo, M Ncube, C Gwanzura, G Bisa, D Gadissa, T Mabuto, L Tsope, I Rabothata, N Ntombela, K Sithole, S Sisay, M Makgabo, RE Chaisson, G Churchyard, CJ Hoffmann, V Chihota

**Affiliations:** The Aurum Institute, Isando, South Africa; Center for Tuberculosis Research, Johns Hopkins University, Maryland, USA; KNCV TB Plus, The Hague, Netherlands; KNCV Ethiopia, Addis Ababa, Ethiopia; Clinton Health Access Initiative, Harare, Zimbabwe; National Department of Health, Pretoria, South Africa; National TB and Leprosy Program, Ministry of Health and Child Care, Harare, Zimbabwe; Addis Ababa Health Bureau, Addis Ababda, Ethiopia; KNCV TB Plus, Addis Ababa, Ethiopia; School of Public Health, University of the Witwatersrand, Johannesburg, South Africa; Department of Medicine, Division of Infectious Diseases, Vanderbilt University, Nashville, TN, USA

**Keywords:** Tuberculosis preventive treatment, people living with HIV (PLWH), routine settings, cascade of care, digital tools

## Abstract

**Background:** TB preventive treatment (TPT) is recommended for those at risk of TB, but despite encouraging trial data, evidence of its effectiveness from routine settings with diverse TPT regimens is limited. We describe routine TPT implementation outcomes in people living with HIV (PLWH) to identify bottlenecks and opportunities for improvement along the cascade of care.

**Methods:** From 2021-2023, we conducted a 24-month prospective cohort study of PLWH in nine health facilities in Ethiopia, South Africa and Zimbabwe. Socio-demographic and clinical data were collected at enrolment. During follow-up, we collected information on TPT initiation, TPT-related side effects and completion. A sub-group of PLWH were allocated digital boxes (evriMED1000) as a proxy measure for assessing treatment completion. TB case ascertainment was based on confirmed diagnoses abstracted from clinic records and symptom agnostic sputum testing initially using Xpert MTB/RIF assay and subsequently MGIT culture at 12- and 24-months post-enrolment.

**Results:** We enrolled 2,095 participants, mostly female (n=1,489; 71.1%), of median age 42 years [interquartile range: 35-50]. Approximately 60% were considered eligible for TPT (n=1,312) and 1,110 (84.6%) were initiated on TPT within 60 days of ART initiation or at a refill visit during the observation period (n=625 on 3HP and n=487 on 6H/12H). TPT completion via digital pillbox assessment was 55.4% (n=406/733). Symptoms compatible with TPT side effects were commonly reported in 3HP vs.6H/12H [190(31.0%) vs. 51(11.0%)]. TB incidence rate was 1.27 (95%CI: 0.95-1.69)/100 person-years during study follow-up in a mostly long-term ART population and was lower for PLWH who completed TPT (aIRR: 0.39; 95% CI: 0.20-0.78, p-value: 0.01).

**Conclusions:** There were losses along the cascade, particularly at completion. In programmatic settings, we observed more potential side effects in 3HP compared to 6H/12H. Interventions to support TPT completion are needed.

**Author Summary:** *What is already known on this topic:*

- Clinical trials have shown that 3HP is well tolerated and has higher completion rates than 6H/12H.
- Identifying bottlenecks to and determinants of successful TPT implementation in programmatic settings is necessary.

*What this study adds:*

- In a multi-site and country cohort of 2,095 PLWH attending routine HIV care, we observed that most individuals could be initiated on TPT and that side effects were generally mild but correlated with not completing TPT
- There are gaps in adhering to guidelines especially on the prescription of TPT to PLWH with prior TB.
- Clinical reporting (defined as date of TPT completion from clinic records) of TPT completion is an overestimation: only slightly more than half of participants initiated on TPT completed it as assessed by the digital pillbox (Medication Event Reminder Monitor-MERM).
- Incident TB disease (often asymptomatic) occurred among 2.5% of participants over the two years of follow-up. TPT, even at the relatively low rates of completion reported by the digital pillbox, appeared to reduce the rate of incident TB disease by 61%.

*How this study might affect research, practice or policy:*

- These findings should reassure clinical providers that most patients can be safely initiated on any TPT regimen, that side effects are generally self-limiting and do not require clinical intervention, and that TPT is effective in protecting against incident TB disease.
- These results emphasize the need for additional approaches to support TPT prescribing and patient-level adherence and regimen completion. Tools to accurately measure TPT completion are required.
- These findings show that providers should be encouraged to initiate TPT to all eligible PLWH to close the existing gaps.
- While improved adherence might lead to a reduced TB incidence rate, the impact with the current programmatic implementation still has public health importance. This is an important point as countries navigate a resource-constrained future.

## Introduction

The 2024 Global Tuberculosis (TB) report estimated that approximately 23% of the global population had TB infection, with about 6% of these individuals progressing to TB disease.^1^ Among individuals with TB disease in 2023, an estimated 662,000 were people living with HIV (PLWH), among whom an estimated 161,000 deaths occurred.^2^ In 2023, according to the 2024 WHO Global Report, overall TB incidence rate per 100,000 in Ethiopia, South Africa and Zimbabwe was 146, 427 and 211 respectively.^2^ The TB incidence rate among PLWH in these countries was 12, 230 and 127 per 100,000 respectively.^2^ There is substantial evidence from studies conducted globally supporting the provision of TB preventive treatment (TPT) using isoniazid to decrease progression to TB disease, reducing risk of TB disease by 40% and risk of death among PLWH.^3, 4^ Based on these observations, TPT is a key TB control strategy recommended by the World Health Organization (WHO).^5^ Though recommended by the WHO since 1991, the delivery of TPT has generally been sub-optimal. Reasons for low uptake include safety concerns, fears of selecting for drug resistance if TB disease is not fully excluded and treatment duration.^6^ While the WHO has recently reported progress in the delivery and uptake of TPT in PLWH,^2^ many settings continue to experience sub-optimal provision and completion of TPT.^6^

The WHO recommends TPT for all PLWH in high TB burden settings who lack evidence of TB disease based on the WHO four symptom screen (any current cough, fever, night sweats, and weight loss) and/or molecular TB testing.^7^ National guidelines in Ethiopia, South Africa and Zimbabwe recommend that all PLWH receive adherence counselling prior to TPT initiation, clinical monitoring for TB disease and laboratory testing for liver toxicity at each ART initiation and/or refill visit.^8–10^ The guidelines also specify the duration of two to six months between each ART refill visit in community-based differentiated service delivery (DSD) models.^8–10^

Recently, shorter course TPT regimens have been recommended, including one-and three-month rifapentine and isoniazid combination regimens,^11–15^ with the three months of weekly rifapentine and isoniazid (3HP) recommended by WHO as the preferred treatment. Successful scale up in routine settings is dependent on identifying and addressing factors related to attrition along the TPT delivery cascade. We sought to describe routine TPT implementation and identify opportunities for improvement by evaluating TPT delivery among PLWH attending routine primary healthcare settings in Ethiopia, South Africa, and Zimbabwe.

## Methods

### Study design and settings

The optimizing TPT delivery among PLWH (Opt4TPT) study was a multisite observational prospective cohort study of PLWH receiving routine HIV care in nine primary healthcare facilities in Ethiopia, South Africa and Zimbabwe. The selected facilities were primary care clinics with established TB programs and had at least 300 monthly HIV/ART clinic encounters and provided 3HP, 6 months of isoniazid (6H), and/or 12 months of isoniazid (12H) to their patients. The expected frequency of monthly ART initiation visits in all nine healthcare facilities was between 39-74 visits. Similarly, 953-7,103 refill visits were expected during the study enrolment period. National guidelines for each country stipulated that all PLWH without TB symptoms, TB or liver disease, or have a TPT history (>2-3 years) or TB treatment should receive TPT.^8–10^

### Sample size considerations

The sample size estimates were based on the outcome of successfully completing 3HP or 6H/12H, and we used data from clinical trials indicating that completion could range from 65% to 97%.^11, 16–19^ We hypothesized that we would observe 90% 3HP, and 65% 6H/12H completion rates in our setting. We aimed to enrol 2,000 PLWH for the cohort study and that we would be able to monitor at least 1,500 PLWH for completion assuming that they were either initiating or refilling ART. We considered a conservative estimate of 15% loss to follow-up which would enable us to estimate completion with a margin of ±3% percentage points for completion rates between 65% and 95%, on a 2-sided 5% level of significance. This sample size was also sufficient to provide estimates of side effects within ±2% percentage points margin of error, if the rates of side effects ranged from 2% to 7% as observed in prior studies.^11, 16–19^

### Study population

The Opt4TPT study was conducted among PLWH attending routine HIV care. TPT assessment, prescription, and monitoring were conducted by healthcare workers as part of routine care. From July-December 2021, healthcare workers referred potential participants attending routine HIV care visits to research staff in the selected healthcare facilities for screening and enrolment into the study. According to the study eligibility criteria, we enrolled adult (≥18 years) participants newly initiated on antiretroviral treatment (ART) or refilling ART in the study clinics regardless of their TPT history and followed them up for 24 months. Participants also had to be willing and able to participate in telephonic study visits, attend in-person clinic visits at 12- and 24-months post enrolment, and be able to provide written informed consent.

### Study procedures

Face-to-face study visits with study staff at months 3, 6, 12 and 24 were aligned with routine clinic visits for people being newly initiated on ART or returning for a refill. We also conducted telephonic visits at months 1, 2, and 21. At enrolment, we collected self-reported data on socio-demographic characteristics, as well as clinical and medical history. At baseline and during follow-up, we abstracted data on TPT initiation and completion and TB diagnoses from clinic files. TPT initiation and completion were defined as having a documented TPT start and stop date in the clinic files. Participants initiating TPT during the study implementation period were offered an evriMED1000 Medication Event Reminder Monitor (MERM) device to measure the opening of boxes as a proxy for monitoring uptake, with medication reminders switched off, until all devices were allocated. At 12- and 24-months post-enrolment, we collected sputum samples from all available participants for microbiological TB investigations. Positive Xpert MTB/RIF Ultra and MGIT culture tests results confirmed TB diagnosis. Additionally, information on TB symptoms (self-reported), mortality and side effects/symptoms (reported to study staff and abstracted from clinic files) on 3HP and 6H/12H was collected. After study completion, we abstracted the total number of PLWH eligible for and those prescribed TPT during the study period from country-specific electronic health records such as district health information systems.

### Outcome measures

The primary outcome was the proportion of participants who initiated TPT within 60 days of ART initiation or refill visit in routine health facilities. Since screening for TPT eligibility was conducted as part of routine care, for the purposes of this analysis, we considered eligibility for TPT as having no documented evidence of prior TPT in the clinic records. A clinical record of TPT completion was defined as a record of dispensing a full course of the TPT regimen without a notation of premature discontinuation.

The secondary outcomes were: i) the proportion of PLWH completing a TPT dose by clinic record or digital pillbox, ii) the proportion of PLWH missing TPT doses and iii) the proportion of participants on TPT that reported side effects/any symptoms. Exploratory outcomes were i) TB incidence rate during follow-up, ii) occurrences of asymptomatic TB 12- and 24-months after enrolment, and iii) the proportion of deaths among PLWH during the follow-up period. TPT completion by digital pillbox was defined as per the WHO guidelines of 80% of possible doses taken for 6H/12H and 90% of possible doses taken for 3HP, within an additional 33% duration extension to allow for completion.^14^ Following these guidelines, treatment completion for 6H was defined as 146 doses in a maximum of 239 days; for 12H, 292 doses within 476 days; and for 3HP, 11 doses in a maximum of 120 days. TPT adherence was measured using the electronic monitoring box. The TPT regimen was defined as the regimen recorded in the evriMED1000 database and the first intake date was used as the TPT initiation date. One dose was counted for each calendar day that the device was opened for 6H/12H while a dose was counted as valid only if it was at least 4 days after the previous dose for 3HP. The symptoms were either reported to study staff during a study visit or to health care staff during routine clinical care.

### Statistical analysis

For continuous outcomes we summarized study cohort characteristics and outcomes using medians and interquartile ranges (IQR). We described categorical variables using frequencies and proportions. Bar graphs were used to summarize the reported TPT-related symptoms and the TPT cascade. Cases with missing values were included in the denominators of categorical variables. We present primary, secondary and exploratory outcome results as proportions. Poisson regression models were employed to summarize and compare all-cause mortality rates between TPT regimens and evaluate predictors of TPT initiation, completion by MERM and TB incidence, and results are presented as risk ratios with corresponding 95% confidence intervals (CI). Variables with a p-value <0.20 at univariate level were included in the multivariate regression models but some variables considered important were a priori included regardless of the p-value. TB incidence rate was calculated to assess the risk of TB over the follow-up period. Sensitivity analyses were conducted to examine the robustness of parameter estimates in the case of missing data. Statistical significance was determined at a two-sided 5% threshold. We conducted all analyses in Stata 17.^20^

## Results

During the enrolment period more than 9,613 PLWH made one or more visits to a study clinic. Of these 721 (7.5%) visits were for initiating ART while 8,892 (92.5%) people were returning for/established on ART care (>6 months on ART). Of the total patient population in the study clinics, 2,263 (23.5%; 754 in Ethiopia, 753 in South Africa, and 756 in Zimbabwe) were referred to study staff for study enrolment. Of those, 2,095 (93%) were eligible for study participation and consented to participate (Figure 1).

**Figure 1:**
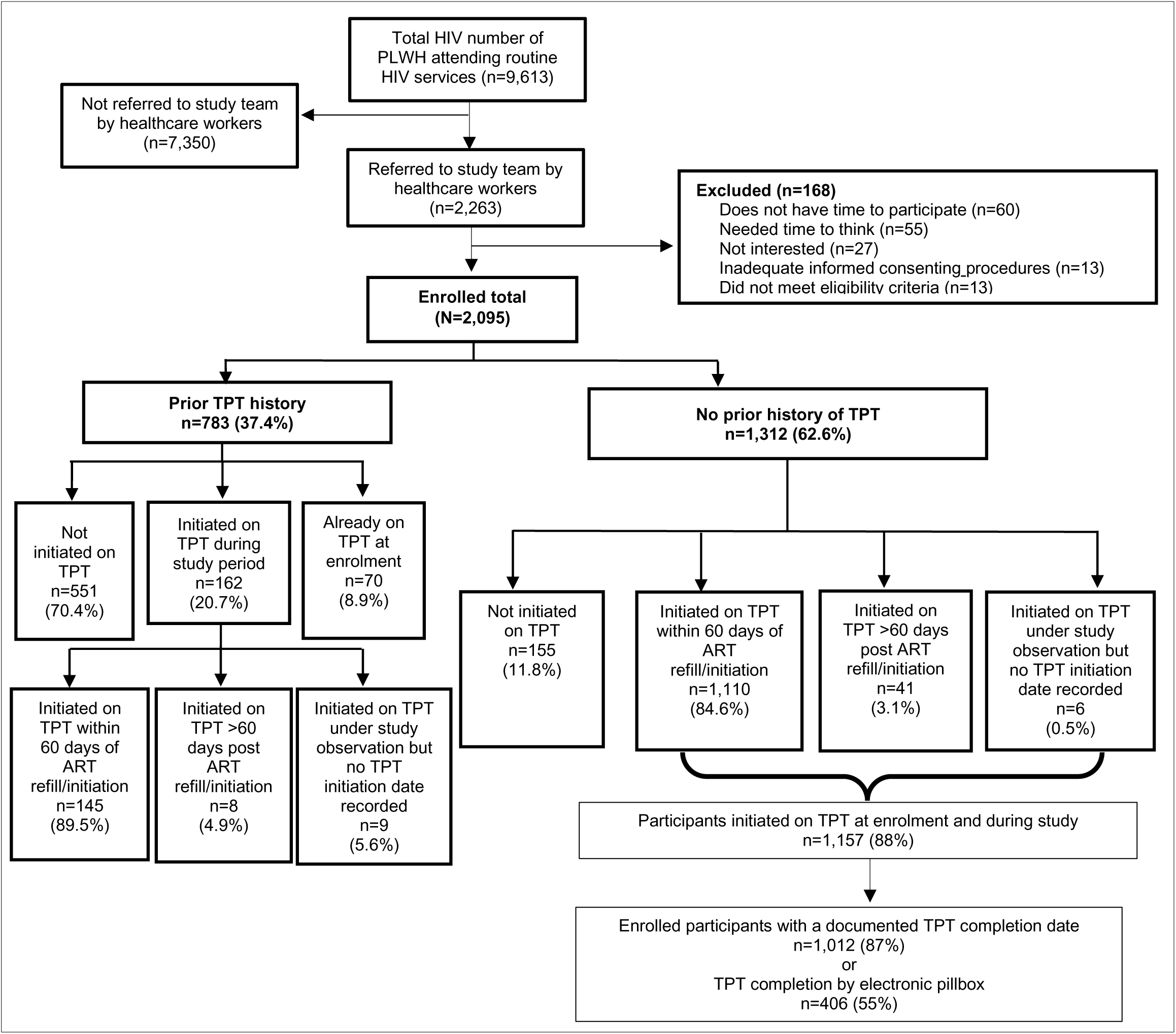
Opt4TPT study flow chart.

Of the 2,095 participants enrolled, most were women (n=1,489, 71.1%), the median age was 42 years (IQR: 35-50), and half reported food insecurity (n=1,098, 52.4%) (Table 1). The majority were established ART patients (n= 1,828, 87.3%). Among those on established ART, 284 (15.5%) reported receiving ART through community-based differentiated service delivery (DSD) models. Nearly 14% of the participants reported a prior TB disease diagnosis (n=287) (Table 1) with notable variation by country: Ethiopia 21.4% (n=143), South Africa 7.2% (n=53) and Zimbabwe 13.1% (n=91), (p<0.001). TPT prior to enrolment was reported by 783 (37.4%) participants; most with prior TPT were in Ethiopia (n=535, 68.3%).

**Table 1:**
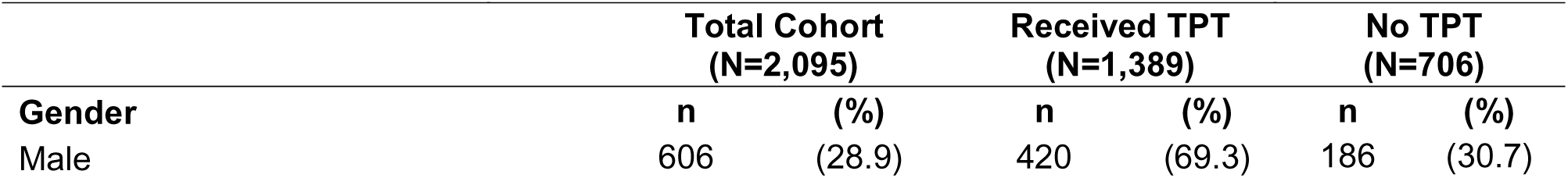
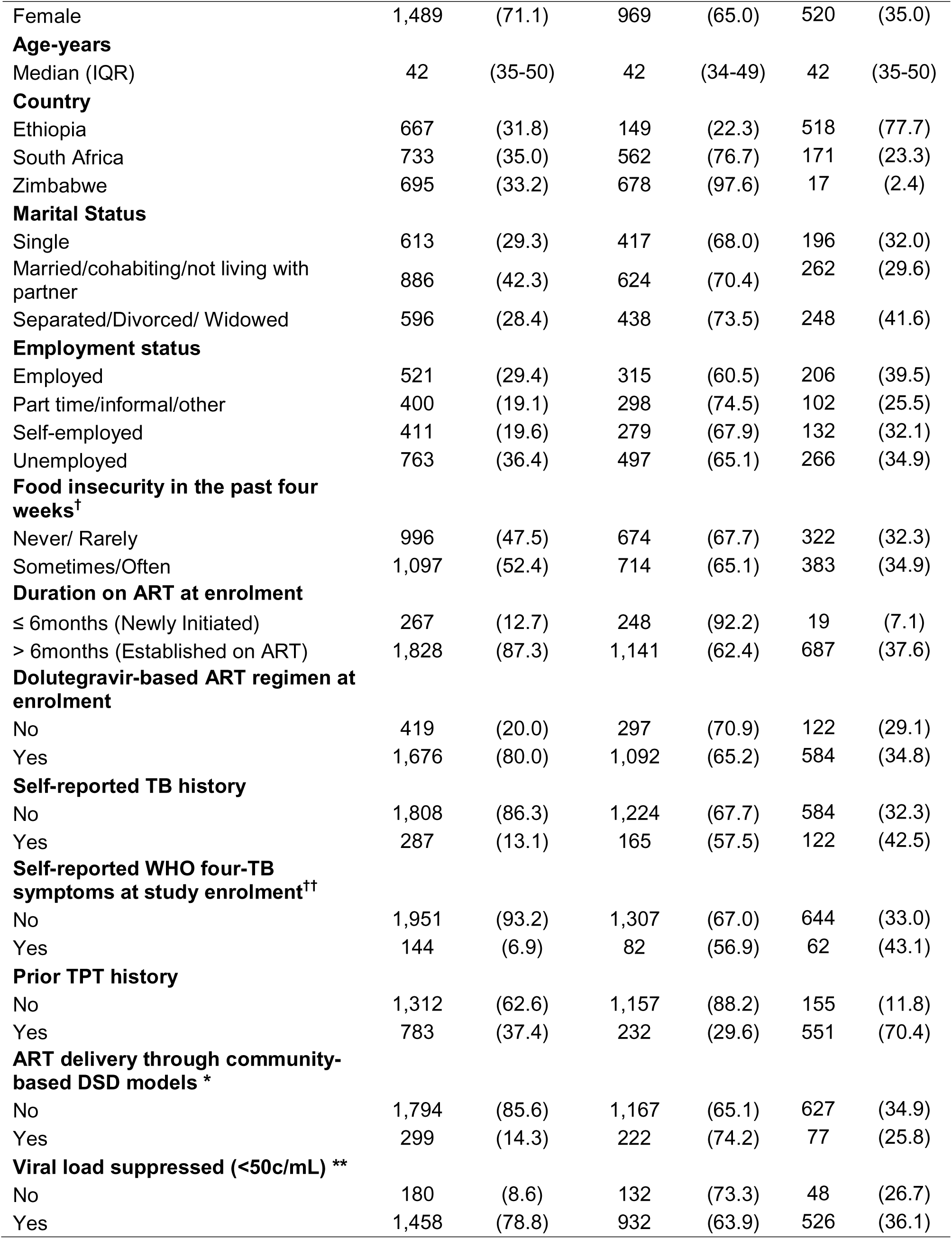
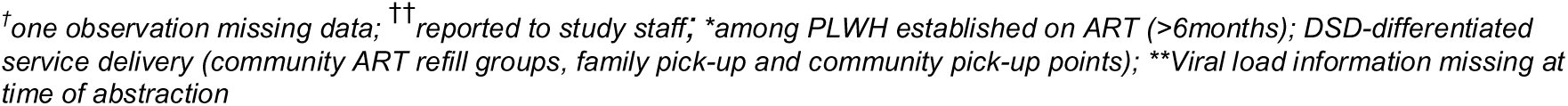
Baseline demographic and clinical characteristics of participating PLWH.

|  | Total Cohort<br>(N=2,095) |  | Received TPT<br>(N=1,389) |  | No TPT<br>(N=706) |  |
| --- | --- | --- | --- | --- | --- | --- |
| Gender | n | (%) | n | (%) | n | (%) |
| Male | 606 | (28.9) | 420 | (69.3) | 186 | (30.7) |

|  | **Total Cohort**<br>**(N=2,095)** |  | **Received TPT**<br>**(N=1,389)** |  | **No TPT**<br>**(N=706)** |  |
| --- | --- | --- | --- | --- | --- | --- |
| Female | 1,489 | (71.1) | 969 | (65.0) | 520 | (35.0) |
| **Age-years** |  |  |  |  |  |  |
| Median (IQR) | 42 | (35-50) | 42 | (34-49) | 42 | (35-50) |
| **Country** |  |  |  |  |  |  |
| Ethiopia | 667 | (31.8) | 149 | (22.3) | 518 | (77.7) |
| South Africa | 733 | (35.0) | 562 | (76.7) | 171 | (23.3) |
| Zimbabwe | 695 | (33.2) | 678 | (97.6) | 17 | (2.4) |
| **Marital Status** |  |  |  |  |  |  |
| Single | 613 | (29.3) | 417 | (68.0) | 196 | (32.0) |
| Married/cohabiting/not living with partner | 886 | (42.3) | 624 | (70.4) | 262 | (29.6) |
| Separated/Divorced/ Widowed | 596 | (28.4) | 438 | (73.5) | 248 | (41.6) |
| **Employment status** |  |  |  |  |  |  |
| Employed | 521 | (29.4) | 315 | (60.5) | 206 | (39.5) |
| Part time/informal/other | 400 | (19.1) | 298 | (74.5) | 102 | (25.5) |
| Self-employed | 411 | (19.6) | 279 | (67.9) | 132 | (32.1) |
| Unemployed | 763 | (36.4) | 497 | (65.1) | 266 | (34.9) |
| **Food insecurity in the past four weeks<sup>†</sup>** |  |  |  |  |  |  |
| Never/ Rarely | 996 | (47.5) | 674 | (67.7) | 322 | (32.3) |
| Sometimes/Often | 1,097 | (52.4) | 714 | (65.1) | 383 | (34.9) |
| **Duration on ART at enrolment** |  |  |  |  |  |  |
| ≤ 6months (Newly Initiated) | 267 | (12.7) | 248 | (92.2) | 19 | (7.1) |
| > 6months (Established on ART) | 1,828 | (87.3) | 1,141 | (62.4) | 687 | (37.6) |
| **Dolutegravir-based ART regimen at enrolment** |  |  |  |  |  |  |
| No | 419 | (20.0) | 297 | (70.9) | 122 | (29.1) |
| Yes | 1,676 | (80.0) | 1,092 | (65.2) | 584 | (34.8) |
| **Self-reported TB history** |  |  |  |  |  |  |
| No | 1,808 | (86.3) | 1,224 | (67.7) | 584 | (32.3) |
| Yes | 287 | (13.1) | 165 | (57.5) | 122 | (42.5) |
| **Self-reported WHO four-TB symptoms at study enrolment<sup>††</sup>** |  |  |  |  |  |  |
| No | 1,951 | (93.2) | 1,307 | (67.0) | 644 | (33.0) |
| Yes | 144 | (6.9) | 82 | (56.9) | 62 | (43.1) |
| **Prior TPT history** |  |  |  |  |  |  |
| No | 1,312 | (62.6) | 1,157 | (88.2) | 155 | (11.8) |
| Yes | 783 | (37.4) | 232 | (29.6) | 551 | (70.4) |
| **ART delivery through community-based DSD models \*** |  |  |  |  |  |  |
| No | 1,794 | (85.6) | 1,167 | (65.1) | 627 | (34.9) |
| Yes | 299 | (14.3) | 222 | (74.2) | 77 | (25.8) |
| **Viral load suppressed (<50c/mL) \*\*** |  |  |  |  |  |  |
| No | 180 | (8.6) | 132 | (73.3) | 48 | (26.7) |
| Yes | 1,458 | (78.8) | 932 | (63.9) | 526 | (36.1) |

<sup>†</sup>one observation missing data; <sup>††</sup>reported to study staff; \*among PLWH established on ART (>6months); DSD-differentiated service delivery (community ART refill groups, family pick-up and community pick-up points); \*\*Viral load information missing at time of abstraction

### TPT initiation

A total of 1,389/2,095 (63.0%) participants had received TPT either before or during the study period (Figure 1). Of these, 716 (51.5%) were initiated on 3HP and 673 (48.5%) on 6H/12H. Most were initiated at study enrolment (1,078; 77.6%) and 67 (4.8%) within seven days of enrolment. An additional 162 (17.6%) were initiated >7 days post-enrolment while 70(3.3%) were already on TPT at study entry. In Ethiopia, most participants had completed TPT one-to-three years prior to enrolment (n=476/667; 71.4%) and may not be eligible for retreatment according to different local guidelines. Of the 2,095 enrolled, 1,312 had no prior documented TPT history and were considered eligible for TPT. Eighty-eight percent (n=1,157/1,312) received TPT and 12% (n=155) did not receive TPT (Figure 2). The majority received TPT within 60 days of ART initiation or a refill visit (n=1,110; 84.6%). Of participants newly initiating on ART, 92.2% (n=248) initiated TPT. Overall drop-off along the cascade is illustrated in Figure 2 and in Supplementary file 1 for country-specific summaries.

**Figure 2:**
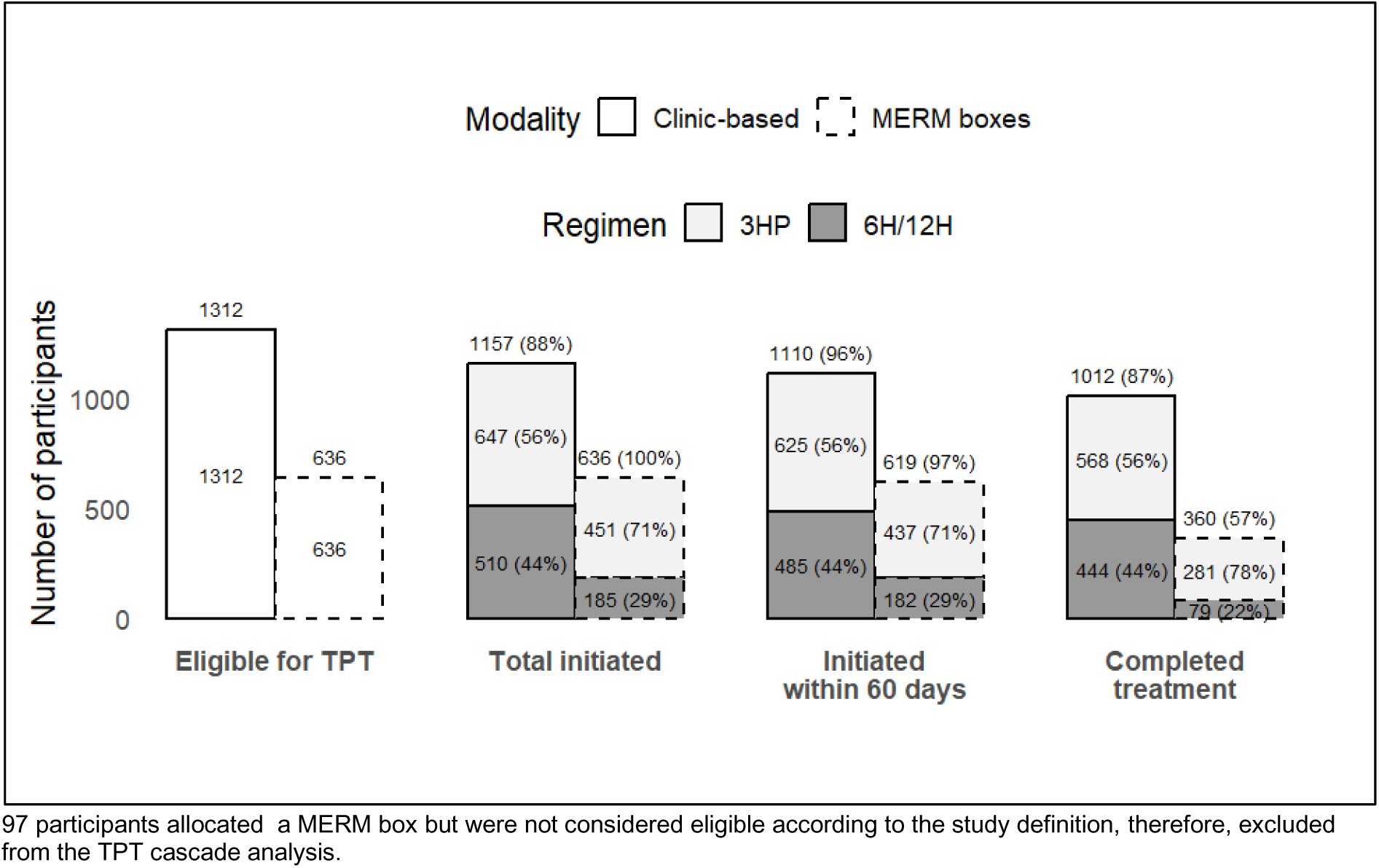
TPT cascade of care, overall, by modality and TPT regimen.

### Factors associated with TPT initiation

TPT was more likely to be initiated among people without prior TPT use (aIRR: 4.0; 95% CI: 2.08-7.69; p-value<0.001) and reported at least one TB symptom (aIRR: 1.82; 95% CI: 1.30-2.78; p-value=0.001). Age, sex, and reporting food insecurity in the past month were not associated with TPT initiation.

### TPT completion among participants Clinic based TPT completion

TPT completion via clinic records showed that 1,012/1,157 (87.5%) participants had a record of full dispensing of medications in the clinic files meeting the clinic-based definition of TPT completion.

### MERM based TPT completion

Of the 880 participants who received TPT and were allocated an electronic pillbox, 733 were included in the MERM analysis. The participant characteristics of PLWH allocated the MERM box are summarized in Supplementary file 2. Exclusions were because of devices never communicated with the server (n=20), the participant was already on TPT at the time the device was allocated (n=22), discrepancies in the TPT prescribing information between the digital pillbox data and the medical record abstractions (n=23), participant reporting withdrawal, death or transfer to another healthcare facility before TPT completion (n=12), or if the participant returned the box prior to TPT completion (n=1). Overall, 406 (55.4%) had MERM evidence of having completed TPT (Figure 2) with 42.1% completion among those who initiated 6H/12H (n=101) and 61.9% completion among those who initiated 3HP (n=305). In adjusted Poisson analysis, participants who initiated 3HP were more likely to complete treatment than those prescribed 6H/12H (aIRR: 1.35; 95% CI: 1.25-1.54), older participants (≥35 years) were more likely to complete treatment than younger participants (aIRR 1.57; 95% CI: 1.25-1.96), those reporting missing at least one TPT dose were less likely to complete TPT (aIRR: 0.77; 95% CI: 0.64-0.94) and individuals with documented TPT completion in clinic records were approximately twice as likely to have completed TPT as assessed by the digital pillbox (aIRR: 2.03; 95% CI: 1.44-2.85) (Table 2). Duration on ART was not associated with TPT completion. Table 2 shows the factors associated with TPT completion. Sensitivity analysis results of 880 PLWH allocated MERM is shown in Supplementary file 3.

**Table 2:** Factors associated with TPT completion among participants assigned digital pillbox (N=733)

| Factor | Crude |  | Adjusted |  |
| --- | --- | --- | --- | --- |
|  | IRR (95%CI) | p-value | aIRR (95%CI) | p-value |
| **Age Category** |  |  |  |  |
| <35 years | 1(Ref) |  | 1(Ref) |  |
| $\geq 35$ years | 1.66 (1.26-2.19) | <0.001 | 1.56 (1.24-1.95) | <0.001 |
| **Sex** |  |  |  |  |
| Male | 1 (Ref) |  |  |  |
| Female | 0.92 (0.75-1.13) | 0.41 | - | - |
| **Duration on ART** |  |  |  |  |
| Newly Initiated | 1 (Ref) |  | 1(Ref) |  |
| Established on ART | 1.23 (0.92-1.74) | 0.14 | 1.02 (0.91-1.15) | 0.73 |
| **TPT Regimen** |  |  |  |  |
| 6H/12H | 1 (Ref) |  | 1(Ref) |  |
| 3HP | 1.48 (1.18-1.85) | 0.001 | 1.36 (1.20-1.51) | <0.001 |
| **Self-reported missed TPT doses** |  |  |  |  |
| No | 1 (Ref) |  | 1(Ref) |  |
| Yes | 0.77 (0.61-0.97) | 0.02 | 0.79 (0.67-0.95) | 0.01 |
| **Receiving ART from a community-based DSD model** |  |  |  |  |
| No | 1 (Ref) |  |  |  |
| Yes | 1.10 (0.85-1.44) | 0.46 | - | - |
| **Reported side effects** |  |  |  |  |
| No | 1 (Ref) |  | 1 (Ref) |  |
| Yes | 1.16 (0.91-1.48) | 0.23 | 1.08 (1.05-1.25) | <0.001 |
| **Reported food insecurity in past four weeks** |  |  |  |  |
| No | 1 (Ref) |  |  |  |
| Yes | 1.05 (0.87-1.28) | 0.61 | - | - |
| **Clinic-based TPT completion** |  |  |  |  |
| No | 1(Ref) |  | 1 (Ref) |  |
| Yes | 2.07 (1.32-3.24) | 0.002 | 2.03 (1.44-2.85) | <0.001 |

### Reported symptoms during follow-up by TPT use

Overall, 360 of the 2,095 (17.2%) participants reported a total of 576 symptoms to the study team: 198 (16.5%) from participants while receiving 6H/12H, 141 (19.1%) from participants while receiving 3HP, and 21 (13.5%) from those not on TPT at any time. Overall, median time (since TPT initiation to reporting these symptoms) was 8.5 (IQR:1-30) days. Symptoms that led to missed TPT doses were reported among 25 (2.2% of all participants receiving TPT). Separately, in file review of symptoms recorded by clinicians, 146 participants reported a total of 161 symptoms. The most common symptoms documented by clinicians were symptoms suggestive of early hepatotoxicity (n=36, 3.8%) among those receiving 3HP and potential hypersensitivity reactions (n=36, 1.3%) among those receiving 6H/12H (Figure 3). No deaths were attributed to medication side effects. Figure 3 summarizes the symptoms reported to both study and clinic staff during follow-up.

**Figure 3:**
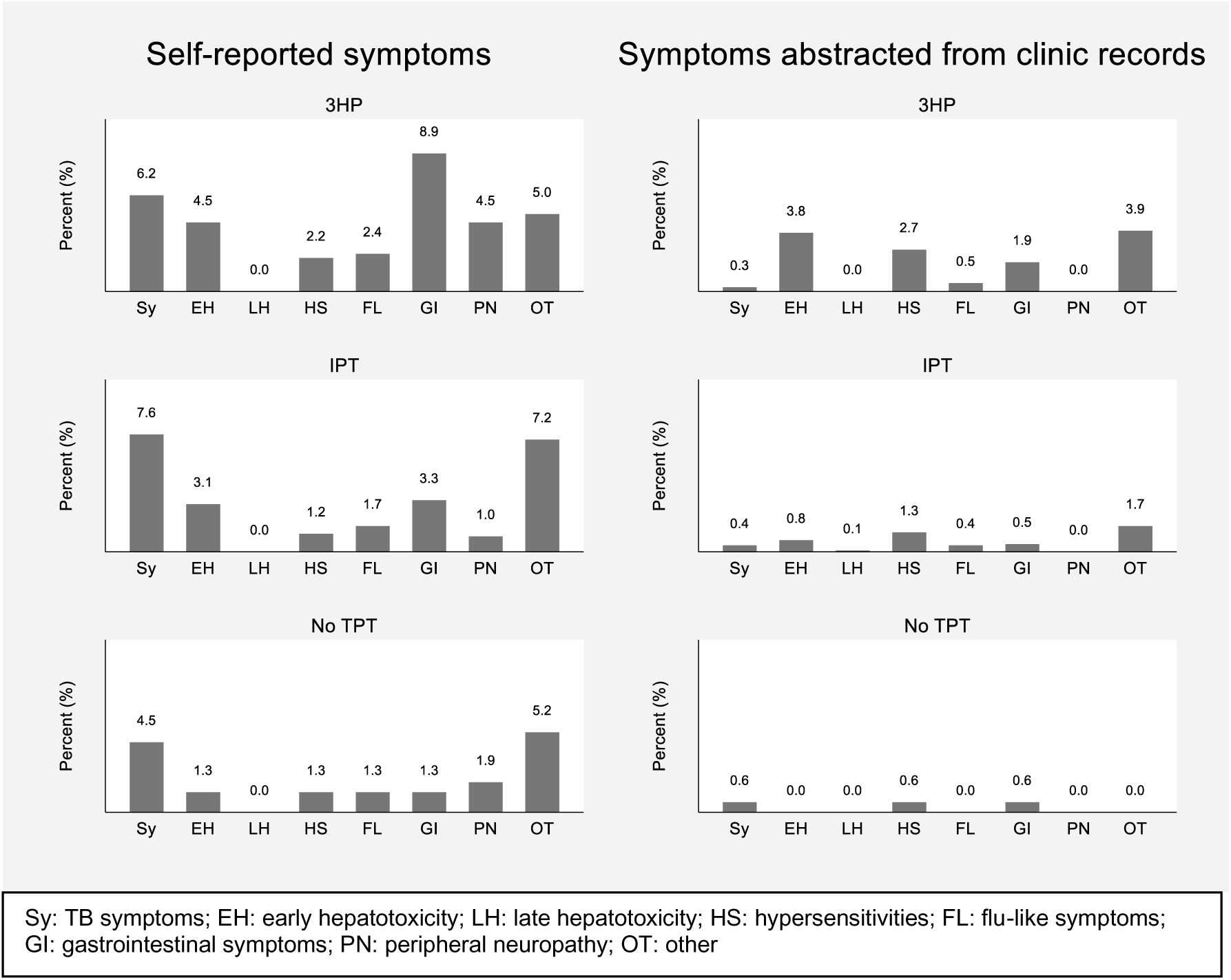
Reported and clinically assessed symptoms by TPT regimen and status.

### Missed TPT doses

Overall, 312 of 1,157 (27%) participants receiving TPT reported missing one or more doses. The most common reasons were forgetting (n=90, 36.0%), time constraints (n=21, 8.4%), medication running out (n=25, 10.0%) and side effects (n=22, 8.8%). More of the participants who reported missed doses due to side effects were receiving 3HP (n=15/110, 13.6%) than 6H/12H (n=7/140, 5.0%). Among participants who completed TPT as assessed via the MERM box, 26.1% (n=204/782) were recorded to have missed at least one TPT dose.

### TB incidence rate during follow-up

During the two years of observation, 51 PLWH were diagnosed with TB. Four were diagnosed within 3 months of enrolment and were considered prevalent disease. Forty-seven were diagnosed between 3 and 24 months after study enrolment either during the 12- or 24-month microbiologic testing or were routinely diagnosed by a healthcare worker at the study clinic. The incidence rate during this period was 1.27 (95% CI: 0.95-1.69) per 100 person years. Among these, 24 (51%) were asymptomatic and detected during the sputum-based microbiological testing 12- or 24-months post-enrolment. The crude TB incidence rate was higher in PLWH established on ART (IRR: 1.47; 95% CI: 0.53-4.10) when compared to those newly initiating. TB disease incidence during the study period was lower among participants who completed TPT (aIRR: 0.39; 95% CI: 0.20-0.78, p-value: 0.01) (Supplementary file 4).

### Mortality rates during follow-up

Twenty-five deaths were recorded of which three were from TB, had no previous TB history and were initiated on 3HP during study observation. Other causes of death were cancer-related (n=6), diabetic complications (n=1), toxoplasmosis (n=1), non-specific infections (n=4), natural causes (n=1), road accident (n=1), and cardiac disease (n=1); seven had no cause of death documented. The all-cause mortality rate was higher in participants not on TPT compared to those on any TPT (Supplementary file 5).

## Discussion

We demonstrated that high levels of TPT initiation are feasible in routine settings: 80% of participants without prior TPT or recent TB treatment received TPT during the study period; 90% among those initiating ART. In addition, TPT was well tolerated and appeared to improve clinical outcomes with a lower incidence rate of TB among those who were initiated on TPT and even lower rate among those who completed TPT. There were no deaths or severe morbid outcomes attributable to TPT. A notable challenge was TPT completion as measured by MERM, with only roughly half of participants completing the regimen with a higher proportion of those who received 3HP compared to 6H or 12H completing.

The proportion of participants who received TPT was higher than national reporting from each country,^2^ but consistent with several recent studies.^11, 13, 21–24^ The study population largely comprised people on established ART. This is programmatically important, as HIV care in many settings is increasingly characterized by a growing number of long-term ART patients. Therefore, these findings characterize the ability to provide TPT to this population, the ongoing high TB disease incidence rate, and the suggestion that TPT (especially completion) reduced TB disease incidence.

Our observed tolerability of TPT, including 3HP, was consistent with clinical trials.^24–26^ Most side effects did not require clinical management or discontinuation of TPT; some led to missed doses. Missing doses due to side effects was reported by a larger proportion of participants on 3HP than on 6H/12H, a finding consistent with a systematic review of TPT adverse events.^27^

We sought to monitor TPT adherence using digital pillboxes to provide a patient-level view of adherence. This is in contrast to the routine clinical practice in the three study countries of reporting completion as having dispensed the full course of a regimen. When operationalized, this often meant that 3HP was documented as completed at the time of initial dispensing because the full regimen was often provided as a single package. Although the digital pillbox also has challenges, the overall completion rate (55%) and higher completion for 3HP (63%) compared to 6H (43%) are plausible and consistent with prior studies.^11, 26, 28^ In addition, our findings show that individuals with documented TPT completion in clinic records were more likely to have completed treatment as assessed by the digital pillbox, suggesting reasonable concordance between routine documentation and objective adherence measures. However, incomplete concordance indicates that clinic records alone may overestimate true completion. We also observed lower incident TB among those who completed TPT suggesting a degree of accuracy for the adherence monitoring. Integrating objective adherence tools could strengthen monitoring and improve confidence in TPT programme outcomes.

We observed a relatively high incidence of TB despite most of our participants being long-term ART patients. This finding is consistent with prior reports of TB incidence of between 0.58 - 4.2 per 100 person years among those on ART.^29, 30^ TB continues to be a burden in the long-term ART population. This reinforces the importance of routine TB testing among those on established ART. In addition, our finding of considerable asymptomatic TB were similar to prior studies globally.^31–34^ Active case finding that is not guided by symptom screening only should be considered an important activity due to the high proportion of asymptomatic disease among people on long-term ART and a considerable proportion of PLWH initiated on TPT despite reporting at least TB symptom.

Our study has the following strengths: Firstly, we demonstrated real-world relevance by reflecting the actual conditions in routine care, highlighting the practical challenges and variations that are not observed in clinical trials. Secondly, the use of diverse settings in southern and eastern Africa with different background rates of TB improves the representativeness. Thirdly, the prospective nature of the study allowed for the tracking of exposures and outcomes supporting causal inference.

### Limitations

Our study has important limitations. Firstly, all participants were referred to study staff by healthcare workers presenting a missed opportunity to objectively assess every potential participant for TPT eligibility. Study-defined TPT eligibility was estimated from the records and defined as having no record of TPT history prior to enrolment. Secondly, healthcare workers likely referred participants for screening into the study when they were prescribing TPT, leading to a biased sample with a high proportion initiating TPT. Notably, our participants represented only a quarter of all the PLWH receiving care from the study facilities. Referral bias may have resulted in a larger proportion of our study participants receiving TPT than the eligible clinic population. Notably, the demographic of our study population roughly reflect those of HIV cohorts in the study countries, suggesting that our participants likely represent the overall ART populations at the study clinics.^35–38^ Implementation of the TB prevention study may have influenced routine practices within these clinics. Thirdly, the electronic monitoring box only recorded whether the device was opened and, thus, is not a direct measurement of medication ingestion. Some participants had greater than 100% calculated adherence which may reflect the use of the device for other storage needs. As such, our estimated completion levels may be an overestimate compared to completion in routine care. Second, the electronic boxes were not large enough to fit a full course of most TPT regimens. Thus, participants were expected to refill the device, as needed. Thus, a participant could have achieved treatment completion without the device recording it they chose not to refill the device. For such participants, our estimated adherence levels would underestimate completion.

## Conclusions

We observed high TPT initiation among people initiating ART and those on long-term ART, a high incidence rate of TB, sub-optimal completion of TPT regimens, and an association between TPT completion and lower TB incidence. Further work is needed to assure high levels of TPT initiation and the development of practical approaches to assessing and supporting adherence to TPT. The high overall burden of TB highlights the importance of TB prevention, including use of TPT with a potential value for repeat courses of TPT after three or more years. TPT remains a critical tool in addressing HIV associated morbidity and mortality among the full spectrum of PLWH including those with advanced HIV disease and those on long-term and effective ART.

## Data availability

The Aurum Institute has an established procedure to make its research data more broadly available. Information on the process for requesting and obtaining data is available on the Aurum Institute website. Datasets used for the analyses of this article, can be requested through an online request lodged on the Aurum website. The request will be assessed by the technical review committee, and once approved the datasets, study protocol and statistical analysis plan will be made available to interested investigators making the request.

## Acknowledgements

The authors are grateful to the administration, staff, and participants at the nine healthcare facilities in Ethiopia, South Africa and Zimbabwe for their time and participation.

## Patient and public involvement

This study was designed without patient participation.

## Ethical approvals

This study was approved by the University of the Witwatersrand Human Research Ethics Committee (No: 210212), Institutional Review Board, Johns Hopkins University (IRB00280677), Medical Research Council of Zimbabwe (MRCZ/A/2727) and Addis Ababa Health Bureau Institutional Review Board (A/A/145/227). We also received approval from the local district research ethics committees in South Africa and permission from the healthcare facilities.

## Author contributions

VC, CJH, GC, RC and JEG conceptualized the study. LC oversaw overall study implementation. NK and AB oversaw the local implementation as site principal investigators. IR, LT, SS, AK, NN and DG supervised data collection. SG and BAN analysed the data. LC wrote the manuscript with input from all authors. CM, KS, JEG, RC, BAN, VC, GC, TM, and AB critically reviewed the manuscript. The authors read and approved the final manuscript.

