## Supplementary File 1-5 for "High initiation but sub-optimal completion of tuberculosis preventive treatment among people living with HIV on ART, persistent tuberculosis incidence and programmatic implications: Findings from a two-year multi-country cohort study"

**Supplementary files**

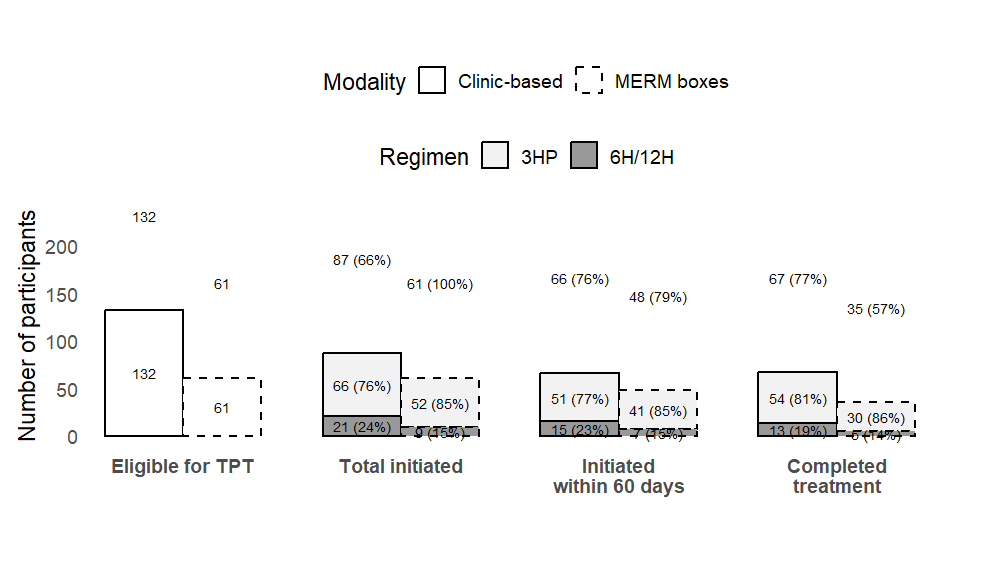

Supplementary file 1a: TPT Cascade of care, Ethiopia

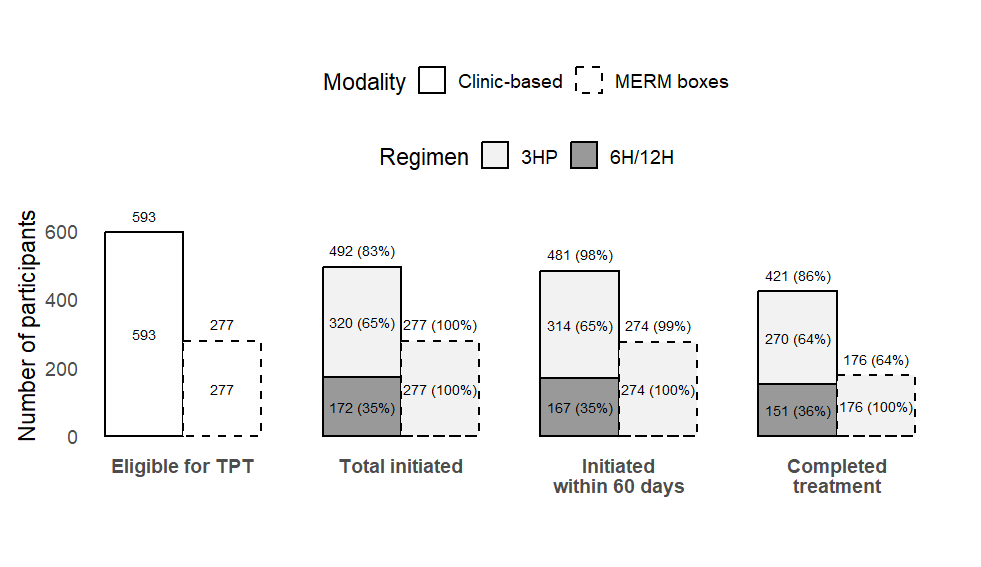

Supplementary file 1b: TPT Cascade of care, South Africa

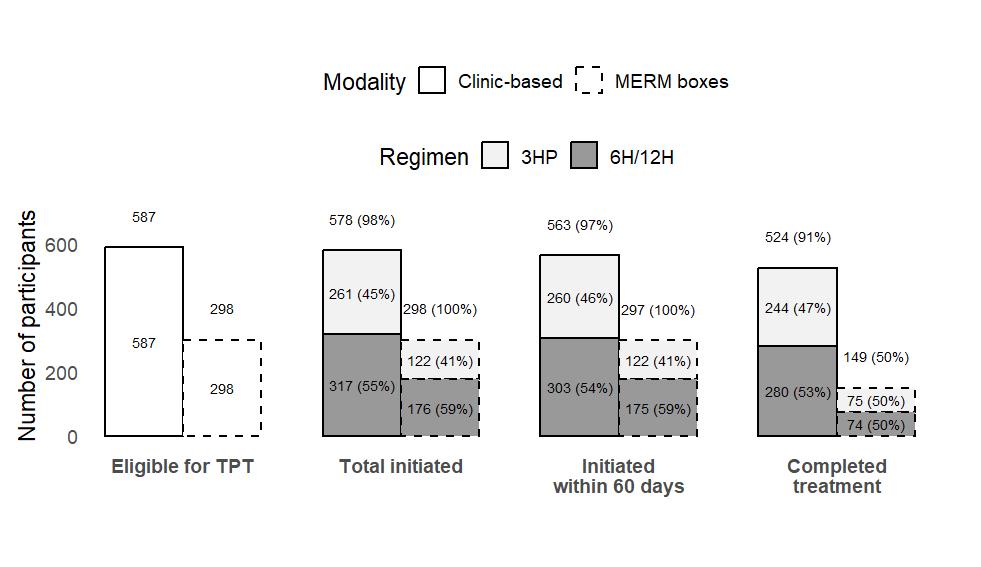
Supplementary file 1c: TPT Cascade of care, Zimbabwe

Supplementary file 2: Participants characteristics by digital pillbox allocation

|  | | Digital pillbox allocation | |
| --- | --- | --- | --- |
|  | Total Cohort | Yes | No |
|  | N=2,095 | N=880 | N=1,215 |
| **Age - Median (IQR)** | 42 (35-50) | 42 (35-50) | 41 (35-50) |
| **Gender** |  |  |  |
| Male | 606 (28.9) | 269 (30.6) | 337 (27.7) |
| Female | 1,489 (71.1) | 611 (69.4) | 878 (72.3) |
| **Country** |  |  |  |
| South Africa | 733 (35.0) | 380 (43.2) | 353 (29.1) |
| Ethiopia | 667 (31.8) | 98 (11.1) | 569 (46.8) |
| Zimbabwe | 695 (33.2) | 402 (45.7) | 293 (24.1) |
| **Marital Status** |  |  |  |
| Single | 613 (29.3) | 263 (29.9) | 350 (28.8) |
| Married/Cohabiting | 886 (42.3) | 386 (43.9) | 500 (41.2) |
| Previously married | 596 (28.4) | 231 (26.3) | 365 (30.0) |
| **Employment Status** |  |  |  |
| Employed | 521 (24.9) | 193 (21.9) | 328 (27.0) |
| Unemployed | 763 (36.4) | 319 (36.3) | 444 (36.5) |
| Informally employed | 400 (19.1) | 206 (23.4) | 194 (16.0) |
| Self-employed | 411 (19.6) | 162 (18.4) | 249 (20.5) |
| **Food Security in past month*** |  |  |  |
| Never/rarely | 996 (47.5) | 417 (47.4) | 579 (47.7) |
| Sometimes/Often | 1,097 (52.4) | 463 (52.6) | 634 (52.2) |
| **ART Status** |  |  |  |
| Newly initiated ART | 267 (12.7) | 153 (17.4) | 114 (9.4) |
| Established on ART | 1,828 (87.3) | 727 (82.6) | 1,101 (90.6) |
| **DTG-based ART regimen** |  |  |  |
| No | 419 (20.0) | 207 (23.5) | 212 (17.4) |
| Yes | 1,676 (80.0) | 673 (76.5) | 1,003 (82.6) |
| **Self-reported TB History** |  |  |  |
| No | 1,808 (86.3) | 774 (88.0) | 1,034 (85.1) |
| Yes | 287 (13.7) | 106 (12.0) | 181 (14.9) |
| **Reported baseline TB symptoms** |  |  |  |
| No | 1,951 (93.1) | 829 (94.2) | 1,122 (92.3) |
| Yes | 144 (6.9) | 51 (5.8) | 93 (7.7) |
| **Receiving ART from a community-based DSD model*** |  |  |  |
| No | 1,794 (85.6) | 756 (85.9) | 1,038 (85.4) |
| Yes | 299 (14.3) | 124 (14.1) | 175 (14.4) |
| **Virally suppressed*** |  |  |  |
| No | 180 (8.6) | 90 (10.2) | 90 (7.4) |
| Yes | 1,458 (69.6) | 572 (65.0) | 886 (72.9) |
| *some observations are missing and column % not adding up to 100% | |  |  |

Supplementary 3: Factors associated with TPT completion among participants assigned a digital pillbox (N=880)

| **Factor** | **Crude** | | **Adjusted** | |
| --- | --- | --- | --- | --- |
|  | IRR (95%CI) | p-value | aIRR (95%CI) | p-value |
| **Age Category** |  |  |  |  |
| <35 years | 1(Ref) |  | 1(Ref) |  |
| ≥35 years | 1.90 (1.45-2.49) | <0.001 | 1.64 (1.38-1.95) | <0.001 |
| **Sex** |  |  |  |  |
| Male | 1 (Ref) |  | 1 (Ref) |  |
| Female | 0.90 (0.74-1.10) | 0.32 | 0.98 (0.90-1.07) | 0.65 |
| **Duration on ART** |  |  |  |  |
| Newly Initiated | 1 (Ref) |  | 1(Ref) |  |
| Established on ART | 1.83 (1.34-2.50) | <0.001 | 1.33 (0.84-2.12) | 0.23 |
| **TPT Regimen** |  |  |  |  |
| 6H/12H | 1 (Ref) |  | 1(Ref) |  |
| 3HP | 1.84 (1.64-2.31) | 0.001 | 1.41 (1.20-1.66) | <0.001 |
| **Self-reported missed TPT doses** | |  |  |  |
| No | 1 (Ref) |  | 1(Ref) |  |
| Yes | 0.78 (0.62-0.98) | 0.03 | 0.81 (0.65-1.01) | 0.07 |
| **Receiving ART from a community-based DSD model** | |  |  |  |
| No | 1 (Ref) |  | 1 (Ref) |  |
| Yes | 1.10 (0.85-1.44) | 0.46 | 0.96 (0.82-1.12) | 0.62 |
| **Reported side effects** |  |  |  |  |
| No | 1 (Ref) |  | 1 (Ref) |  |
| Yes | 1.18 (0.93-1.50) | 0.17 | 1.05 (0.92-1.21) | 0.09 |
| **Reported food insecurity in past four weeks** | |  |  |  |
| No | 1 (Ref) |  | 1 (Ref) |  |
| Yes | 1.03 (0.85-1.25) | 0.75 | 1.06 (0.92-1.21) | 0.43 |
| **Clinic-based TPT completion** | |  |  |  |
| No | 1 (Ref) |  | 1 (Ref) |  |
| Yes | 2.20 (1.45-3.35) | <0.001 | 2.13 (1.60-2.86) | <0.001 |

Supplementary 4: Predictors of incident TB disease (N=869)

| **Factor** | **Crude** | | **Adjusted** | |
| --- | --- | --- | --- | --- |
|  | IRR (95%CI) | p-value | aIRR (95%CI) | p-value |
| **Age Category** |  |  |  |  |
| <35 years | 1(Ref) |  | 1(Ref) |  |
| ≥35 years | 0.47 (0.13-1.67) | 0.24 | 0.41(0.13-1.29) | 0.13 |
| **Sex** |  |  |  |  |
| Male | 1 (Ref) |  | 1 (Ref) |  |
| Female | 0.65 (0.18-2.30) | 0.50 | 0.67(0.34-1.32) | 0.25 |
| **TPT completion by digital pillbox** |  |  |  |  |
| No | 1 (Ref) |  | 1 (Ref) |  |
| Yes | 0.43 (0.11-1.68) | 0.23 | 0.39 (0.20-0.78) | 0.01 |
| **ART Status** |  |  |  |  |
| Newly initiated | 1 (Ref) |  | - |  |
| Established on ART | 1.47 (0.53-4.10) | 0.57 | - | - |

Supplementary 5: All-cause mortality rate over the two-year follow-up period

|  | **n** | **N** | **Person-years (per 100)** | **Mortality rate**  **(per 100) and 95%CI** | **Mortality rate ratio (per 100) (95%CI), p-value** |
| --- | --- | --- | --- | --- | --- |
| Total deaths | 25 | 2,095 | 3,742 | 0.67 (0.45-0.99) | - |
| Any TPT^†^ | 19 | 1,940 | 3,513 | 0.54 (0.35-0.85) | Reference |
| No TPT | 6 | 155 | 230 | 2.61 (1.17-5.82) | 4.83 (1.93-12.1),  p-value<0.001 |

^†^ This refers to all participants who were initiated on TPT before and after study enrolment
